# Trans-ethnic Mendelian randomization study reveals causal relationships between cardio-metabolic factors and chronic kidney disease

**DOI:** 10.1101/2020.09.04.20188284

**Authors:** Jie Zheng, YueMiao Zhang, Humaira Rasheed, Venexia Walker, Yuka Sugawara, JiaChen Li, Yue Leng, Benjamin Elsworth, Robyn E. Wootton, Si Fang, Qian Yang, Stephen Burgess, Philip C. Haycock, Maria Carolina Borges, Yoonsu Cho, Rebecca Carnegie, Amy Howell, Jamie Robinson, Laurent F Thomas, Ben Michael Brumpton, Kristian Hveem, Stein Hallan, Nora Franceschini, Andrew P. Morris, Anna Köttgen, Cristian Pattaro, Matthias Wuttke, Masayuki Yamamoto, Naoki Kashihara, Masato Akiyama, Masahiro Kanai, Koichi Matsuda, Yoichiro Kamatani, Yukinori Okada, Min Xu, YuFang Bi, Guang Ning, George Davey Smith, Sean Barbour, CanQing Yu, Bjørn Olav Åsvold, Hong Zhang, Tom R. Gaunt

## Abstract

**BACKGROUND:** The chronic kidney disease (CKD) public health burden is substantial and has not declined as expected with current interventions on disease treatments. A large number of clinical, biological, and behavioural risk factors have been associated with CKD. However, it is unclear which of them are causal.

**OBJECTIVE:** To systematically test whether previously reported risk factors for CKD are causally related to the disease in European and East Asian ancestries.

**DESIGN:** Two-sample Mendelian randomization (MR) and non-linear MR analyses.

**PARTICIPANTS:** 53,703 CKD cases and 960,624 controls of European ancestry from CKDGen, UK Biobank and HUNT, and 13,480 CKD cases and 238,118 controls of East Asian ancestry from Biobank Japan, China Kadoorie Biobank and Japan-Kidney-Biobank/ToMMo.

**MEASURES:** Systematic literature mining of PubMed studies identified 45 clinical risk factors and biomarkers with robustly associated genetic variants, including phenotypes related to blood pressure, diabetes, glucose, insulin, lipids, obesity, smoking, sleep disorders, nephrolithiasis, uric acid, coronary artery disease, bone mineral density, homocysteine, C-reactive protein, micro-nutrients and thyroid function, which were selected as exposures. The outcome was CKD (defined by clinical diagnosis or by estimated glomerular filtration rate (eGFR) < 60 ml/min/1.73m^2^).

**RESULTS:** Eight risk factors showed evidence of causal effects on CKD in European ancestry, including body mass index (BMI), hypertension, systolic blood pressure, high density lipoprotein cholesterol, apolipoprotein A-I, lipoprotein A, type 2 diabetes (T2D) and nephrolithiasis. In East Asian ancestry, BMI, T2D and nephrolithiasis showed evidence of causal effects on CKD. Hypertension showed reliable evidence of a strong causal effect on CKD in Europeans but in contrast appeared to show a null effect in East Asians, suggesting the possibility of different causal risk factors in Europeans and East Asians. Although liability to T2D showed consistent effects on CKD, the effect of glycemic traits on CKD was weak, suggesting T2D may have glucose-independent mechanisms to influence CKD. Nonlinear MR indicated a threshold relationship between genetically predicted BMI and CKD, with increased risk at BMI above 25 kg/m^2^.

**LIMITATION:** Due to the unbalanced distribution of data between ancestries, we could only test 17 of the 45 risk factors in East Asian participants.

**CONCLUSIONS:** Eight CKD-associated risk factors showed evidence of causal effects on the disease in over 1.2 million European and East Asian ancestries. These risk factors were predominantly related to cardio-metabolic health, which supports the shared causal link between cardio-metabolic health and kidney function. This study provides evidence of potential intervention targets for primary prevention of CKD, which could help reduce the global burden of CKD and its cardio-metabolic co-morbidities.

**Research in context:** *Evidence before this study:* Chronic kidney disease (CKD) has a major effect on global health, both as a direct cause of morbidity and mortality, and as an important complication for cardio-metabolic diseases. However, even with the existing interventions, the burden of CKD has not declined as expected over the last 30 years. Existing epidemiological studies of CKD have mainly focused on disease treatment in patients from specific populations and estimated association rather than causality. A systematic assessment of the causal determinants of CKD in different populations is urgently needed, to help promote a shift from treatment of CKD patients to prevention of the disease in high-risk groups. The use of genetic data and the latest Mendelian randomization (MR) methodologies offers a cost-effective way to evaluate the potential intervention targets for prevention of CKD in high-risk groups.

*Added value of this study:* In this study, we systematically constructed a causal atlas of 45 risk factors on CKD in European and East Asian ancestries using MR. To maximise power of these analyses and accuracy of the findings, we collected and harmonised CKD genetic association data from six large-scale biobanks (in over 1.1 million Europeans and 250,000 East Asians). By applying a comprehensive MR framework, including linear two-sample MR, bidirectional MR, multivariable MR and non-linear MR approaches, we identified eight risk factors with reliable evidence of causal effects on CKD in European ancestry studies, including body mass index (BMI), hypertension, systolic blood pressure, high density lipoprotein cholesterol, apolipoprotein A-I, lipoprotein A, type 2 diabetes (T2D) and nephrolithiasis. In East Asian studies, BMI, T2D and nephrolithiasis also showed causal effects on CKD. Among other factors, hypertension showed reliable evidence of a strong causal effect on CKD in Europeans but in contrast appeared to show a null effect in East Asians. This MR finding together with previous literature evidence opens up the possibility that hypertension could play different causal roles on CKD across ancestries. For diabetes and glycemic phenotypes, our MR and sensitivity analyses suggested the causal role of liability of T2D on CKD but suggested weak effects of glycemic phenotypes on CKD. This aligns with the recent trial of SGLT2 inhibitors on kidney disease, which implies T2D may have glucose-independent mechanisms to influence CKD. For lipids phenotypes, we found good evidence to support the role of high-density lipoprotein cholesterol on CKD and further suggested the effects of two lipids targets: circulating CETP level and lipoprotein A concentration. For body weight, our study quantified a threshold relationship between BMI and CKD, with increased risk at BMI above 25 kg/m^2^. The causal relationship between nephrolithiasis and CKD were reported in previous studies, but our study confirmed the causal links between the two for the first time.

*Implication of all the available evidence:* This study makes a significant advance in comprehensively prioritising intervention targets for CKD in over 1.2 million participants. Our study presents causal evidence from both European and East Asian population samples, widening the generalisability of the causal atlas. Importantly, the prioritised risk factors are predominantly related to cardio-metabolic health, which supports the shared causal link between cardio-metabolic health and kidney function. Clinically, the high-quality evidence from this study highlights the value of exploring these causal factors in the general population and prioritizes drug targets and life-style interventions for CKD primary prevention, which could help reduce the global burden of CKD and its cardio-metabolic co-morbidities.

---

Chronic kidney disease (CKD) affects 10-15% of the population worldwide. It has a major effect on global health, both as a direct cause of morbidity and mortality, and as an important complication for cardio-metabolic diseases (1)(2)(3). From 1990 to 2017, the global age-standardized mortality for many important non-communicable diseases, such as cardiovascular disease, declined by 30.4%, but till now the mortality decline for CKD was only 2.8% (4). Therefore, reducing CKD incidence could be the next intervention target to reduce the burden of both CKD and its associated-complications globally (5). However, in reality, disease awareness of CKD is still deficient among the general public and health-care authorities. The majority of intervention trials have focused on disease treatment rather than primary prevention of CKD. In the literature, impaired fasting glucose, high blood pressure and high body-mass index are among the leading risk factors for CKD. However, even with the interventions on controlling glucose and blood pressure, the burden of CKD has not declined as expected (4). Thus, a systematic assessment of the causal determinants of CKD is urgently needed, which will help promote a shift from treatment of CKD patients to prevention of the disease in high-risk groups.

Well-designed and well-powered randomized controlled trials (RCTs) are usually the best approach to estimate a causal relationship between a risk factor and a disease. Currently, most published CKD studies were observational and/or RCTs in CKD patients, and these have identified risk factors for CKD progression. However, no reliable RCT evidence exists to support their causal roles on CKD incidence. Mendelian randomization (MR) is an epidemiological method that can be used to obtain evidence about the effect of modifying intervention targets (6). MR exploits the random allocation of genetic variants at conception and is therefore less susceptible to confounding and reverse causality than traditional observational studies. The increasing availability of genetic association resources provides a timely opportunity to test the causal effects of various risk factors on CKD and provide insights into the disease pathogenesis using MR (7)(8).

In this study, we aimed to investigate the causal effects of 45 previously reported risk factors on CKD in general European and East Asian populations. To achieve this, a systematic search of risk factors for CKD was conducted in PubMed using the literature mining tool MELODI (9)(10). A set of two-sample linear MR analyses was performed using CKD and estimated glomerular filtration rate (eGFR) summary data from over 1 million participants from CKDGen consortium (11), UK Biobank (12), Nord-Trøndelag Health (HUNT) Study (13), Biobank Japan (14), China Kadoorie Biobank (15) and Japan-Kidney-Biobank/ToMMo consortium, in conjunction with the largest available genome-wide association study (GWAS) for risk factors in European and East Asian ancestries. A set of comprehensive follow-up analyses was carried out to understand the independent effect of key risk factors and the relationship between glycemic, blood pressure and lipid phenotypes on CKD. For key causal risk factors with MR evidence, non-linear MR (16)(17)(18) was performed in participants from UK Biobank and HUNT Study to investigate the optimal level of intervention.

## METHODS

### Study design

Our study consisted of four components (**Figure 1**). First, we identified 45 risk factors for CKD by mining the literatures in PubMed. Second, we estimated the causal effects of these risk factors on CKD prevalence and eGFR in CKDGen (11), UK Biobank (12), HUNT Study (13), Biobank Japan (14) and China Kadoorie Biobank (15) and Japan-Kidney-Biobank/ToMMo consortium separately using two-sample linear MR approach. We further evaluated the MR evidence based on the strength and consistency of MR evidence across MR methods and across studies and fitness of MR assumptions. Third, we conducted extensive follow-up analyses to confirm the findings for four types of cardio-metabolic risk factors: blood pressure phenotypes, BMI, blood lipid phenotypes and glycemic phenotypes on CKD. Finally, non-linear MR was performed to estimate the optimal level of BMI and fasting glucose for reducing CKD risk in UK Biobank and HUNT Study.

**Figure 1.**
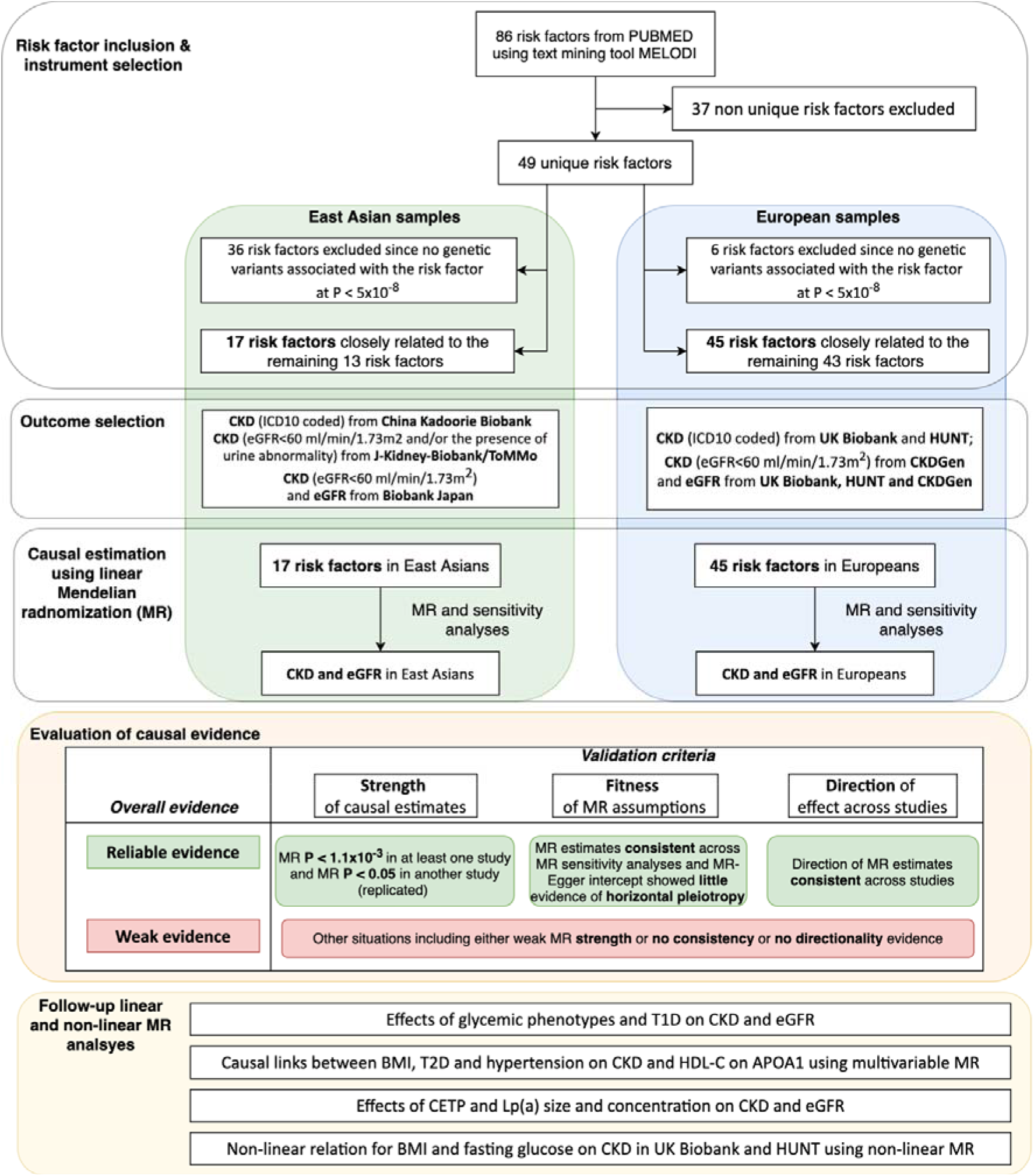
Study design of the trans-ethnic Mendelian randomization study of chronic kidney disease.

### Selection of risk factors

CKD risk factors were identified from a literature review using the MELODI-Presto (9)(10) to search the PubMed database (for more details on this method see **Supplementary Note 1**). We identified 49 risk factors/phenotypes for CKD, including blood pressure phenotypes, diabetic phenotypes (glucose- and insulin-related phenotypes), lipids phenotypes, obesity, smoking, alcohol intake, sleep disorders, nephrolithiasis, serum uric acid, coronary artery disease, bone mineral density, homocysteine, C-reactive protein, micro-nutrient phenotypes (serum metals and vitamins), dehydration and thyroid phenotypes, were selected. By searching the largest available GWAS studies (but minimums the sample overlap with the outcome samples), we extracted genetic variants associated with 45 of 49 risk factors in European ancestry studies and with 17 of 49 risk factors in East Asian ancestry studies and used them as genetic instruments for the MR analyses (**Supplementary Table 1**, more details in **Supplementary Note 1**). To select the independent genetic variants, the genome wide significant SNPs were grouped by LD (r^2^< 0.001 for SNPs within 1Mb genomic region) and only the SNP with the lowest P value per group was kept (**Supplementary Table 2, 3**).

### Association of genetic variants with CKD and eGFR

CKD was defined by clinical diagnosis (International Classification of Diseases (ICD) 10th Revision) in the UK Biobank (12) (the CKD cases were defined as participants with ICD10 code range from N17-N19; the participants with all types of kidney disorders [N00-N29] were excluded from the controls to reduce the possibility of including CKD case in the control group), HUNT (13) and China Kadoorie Biobank (15); defined as eGFR < 60 ml/min/1.73m^2^ in CKDGen (11) and Biobank Japan (14) and defined as eGFR< 60 ml/min/1.73m^2^ and/or the presence of urine abnormality in the Japan-Kidney-Biobank/ToMMo consortium, where eGFR was estimated from serum creatinine using the Chronic Kidney Disease Epidemiology Collaboration (CKD-EPI) formula (19). The genetic associations with CKD and eGFR were reported in three studies of European ancestry (CKDGen: 41,395 cases, 439,303 controls, 8.7% diabetes patients; UK Biobank: 9,884 cases, 454,323 controls, 5.2% with diabetes; and HUNT Study: 2,424 cases, 66,998 controls, 4.9% with diabetes). The genetic associations with CKD were reported in three East Asian studies (Biobank Japan: 8,586 cases, 133,808 controls, 10.2% with diabetes; China Kadoorie Biobank: 848 cases, 94,887 controls, 6.7% with diabetes; Japan-Kidney-Biobank/ToMMo consortium: 4,046 cases, 9,423 controls, 7.3% with diabetes) and eGFR were reported in Biobank Japan (**Supplementary Table 4, Supplementary Note 2**). The novel GWAS results for eGFR and CKD generated using UK Biobank data were accessed using MRC-IEU OpenGWAS database (https://gwas.mrcieu.ac.uk/) (20) and MR-Base (http://www.mrbase.org) (21).

### Statistical analysis

MR is an instrumental variable method that uses genetic variants as instruments to test the causal relationships between an exposure (e.g. BMI) and outcome (e.g. CKD). The causal effect of the exposure and the outcome can be estimated when the three core assumptions are satisfied (**Supplementary Figure 1** and **Supplementary Note 3**). For the binary exposures (e.g. T2D), the odds ratios (ORs) were converted (multiplying log(ORs) by log(2) (equal to 0.693) and then exponentiating) in order to represent the OR of outcome per doubling of the odds of susceptibility to the exposure (22)(23).

A set of two-sample MR and sensitivity analyses for testing the underlying assumptions, were conducted using the TwoSampleMR package (21), including Wald ratio (24), inverse variance weighted (IVW) MR (25), MR-Egger (26), MR weighted median (27), MR mode estimator (28), Steiger filtering (29) and heterogeneity test across multiple instruments (30). Information of the bidirectional MR (31) of genetic liability of CKD or eGFR as exposure and risk factors as outcomes, as well as multivariable MR (MVMR) analyses of correlated phenotypes can be found in Supplementary Note 4. A conservative Bonferroni corrected threshold (α = 1.11×10^−3^, as 45 risk factors were assessed) was used to account for multiple testing. Power calculations were performed using an online tool (https://sb452.shinyapps.io/power/) (32).

### Follow-up MR analyses

We conducted extensive follow-up analyses to validate and extend the depth of our main findings. First, a set of analyses was conducted to understand the pathways linking T2D with CKD: 1) we validated the causal effects of the eight glycemic phenotypes on CKD using Steiger filtering (29) and radial MR (33); 2) we considered the influence of genetic liability of type 1 diabetes (T1D) (34) (**Supplementary Table 2**) on CKD; 3) participants with eGFR measurements were stratified into diabetic (N = 11,529) and non-diabetic populations (N = 118,460) (35) and we conducted MR analyses of T2D and 5 glycemic phenotypes on eGFR in these two sub-populations; 4) diabetic retinopathy was included as a positive control outcome to validate the analytical approach. The instruments for T2D and glycemic phenotypes were used as exposure, while the CKD data from CKDGen, UK Biobank and HUNT as well as the diabetic retinopathy data from UK Biobank SAIGE release (36) were used as outcomes (**Supplementary Table 4**).

Second, to further validate the MR findings of lipids on CKD, the following analyses were conducted: 1) we tested the independent effect of high density lipoprotein cholesterol (HDL-C) and apolipoprotein A-I on CKD using a MVMR model (**Supplementary Note 4**); 2) we estimated the effect of circulating cholesteryl ester transfer protein (CETP) levels (37) on CKD (**Supplementary Table 2**); 3) given that lipoprotein A (Lp[a]) levels for a fixed apo(a) isoform size may vary, we estimated the effect of apo(a) isoform size on CKD (Lp[a] KIV2 repeats and apolipoprotein[a] protein isoform size data from Saleheen et al. (38)) (**Supplementary Table 2**).

Third, to validate the blood pressure and HDL-C MR results in East Asians, we conducted two sensitivity analysis: 1) given the unbalanced distribution between the European and East Asian GWAS resources (39), we need to consider the potential influence of unbalanced number of instruments and the power of the MR results across the two populations. Therefore, we took all SNPs that are associated with systolic blood pressure (SBP; 197 SNPs), diastolic blood pressure (DBP; 235 SNPs) and HDL-C (145 SNPs) in Europeans and checked whether their genetic associations replicated in East Asians GWASs (which the P value thresholds of 0.001 and 0.01 were used as the threshold of replication separately) and then included those replicated SNPs in the MR analyses of SBP, DBP and HDL-C (**Supplementary Table 5**; 128 SNPs were included using P< 0.01, 84 SNPs were included using P< 0.001) on CKD in East Asian samples. 2) We extracted genetic instruments of blood pressure phenotypes from a recent Chinese study (40) and conducted a validation of MR findings between the four blood pressure phenotypes (including hypertension, SBP, DBP, pulse pressure [PP]) and CKD in the three CKD outcome studies using East Asian samples.

Finally, for BMI and fasting glucose, a fractional polynomial approach (17)(18) was applied to estimate the non-linear shape of the association between these risk factors and CKD using UK Biobank and HUNT Study (**Supplementary Note 5**).

### Evaluation of Mendelian randomization evidence

Previous study suggested that P value threshold should not be the only criteria to define “significance” (41). We therefore evaluated the MR evidence using three criteria: (1) **MR strength**: whether the MR IVW estimate of each risk factor passed the Bonferroni-corrected P value threshold (P < 1.1 × 10^−3^) in one study and passed the replication threshold (P < 0.05) in at least one other study; (2) **Fitness of MR assumptions:** whether the MR estimates of each risk factor showed the same direction of effect across MR sensitivity analyses (e.g. MR IVW, MR-Egger and other methods) and checked the influence of horizontal pleiotropy using MR-Egger intercept term and heterogeneity test (Cochran Q for IVW and Rucker’s Q for MR-Egger). (3) whether the **direction of MR effect** of each risk factor on CKD was consistent across multiple studies. **Figure 1** demonstrated how the MR evidence were determined in Europeans and East Asians separately: “Reliable evidence” refers to risk factors that fulfilled all the three criteria and “Weak evidence” refers to risk factors that do not align with any one of the three criteria (e.g.MR estimates with strong MR evidence but with inconsistent directionality).

## RESULTS

### Causal effects of risk factors on CKD

The causal effects of the 45 risk factors on CKD in Europeans and 17 risk factors in East Asians are presented in **Figure 2**. Among them, eight risk factors showed reliable evidence, and the remaining 37 showed weak evidence of associations with CKD. Detailed evaluations of the causal evidence in Europeans and East Asians were presented in **Supplementary Table 6A** and **6B separately**.

**Figure 2.**
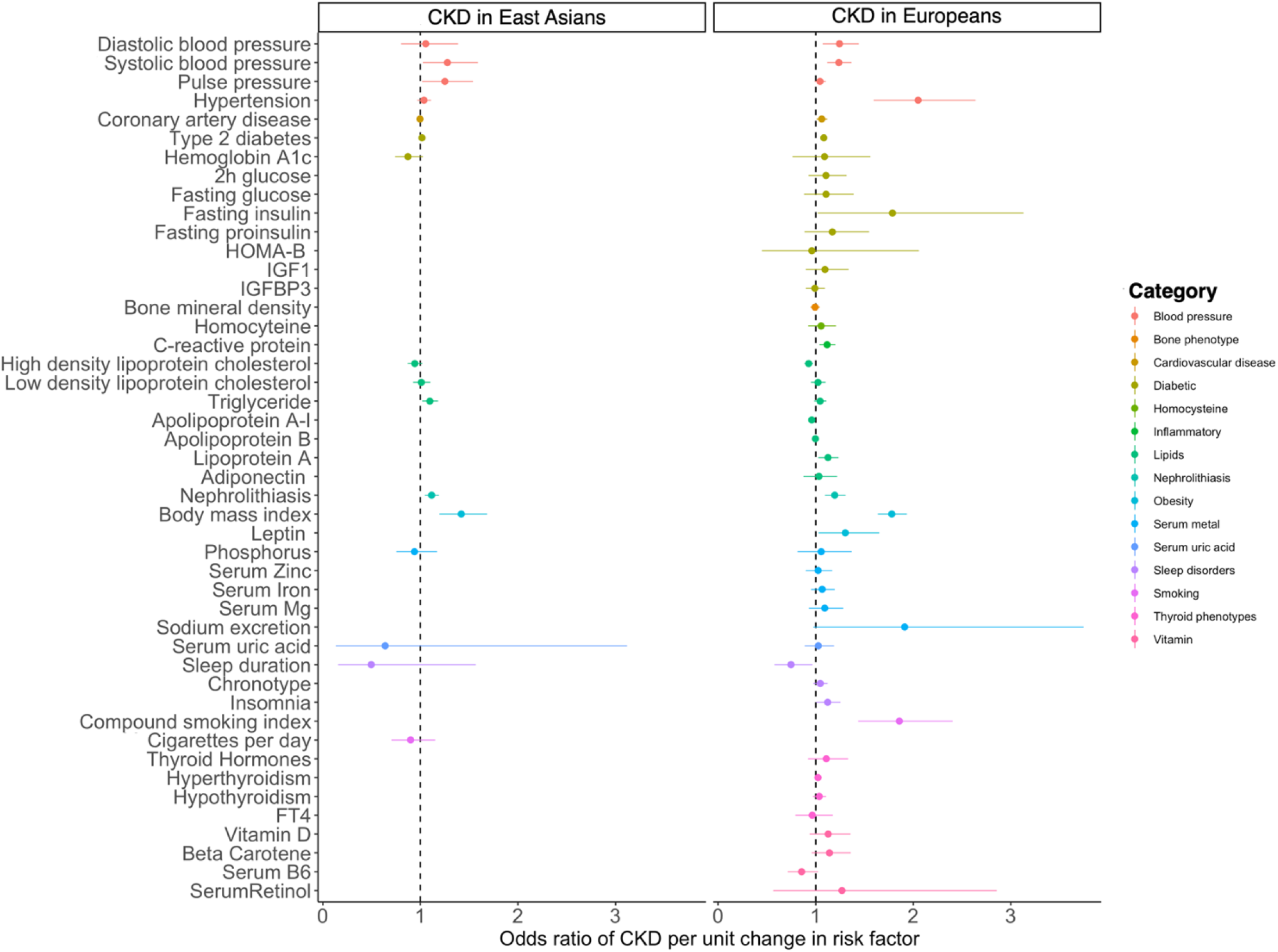
Forest plot for causal effects of the 45 risk factors on CKD in European and Eastern Asians. A) causal estimates using Europeans data; B) causal estimates using Eastern Asians data. For binary exposures, the effect reported on the X-axis is the OR of CKD per doubling in the odds of the exposure. For continuous exposure, the effects on the X-axis is the OR of CKD per 1 SD change in the exposure.

#### Risk factors showing reliable MR evidence

**Supplementary Figure 2A** shows the genetic predictors of eight risk factor are associated with CKD in European ancestries. The IVW odds ratio (95% CIs) for CKD per 1-SD increase in continuous risk factors was 1.78 (1.64 to 1.94) for BMI, 1.24 (1.12 to 1.37) for SBP, 1.13 (1.07 to 1.19) for Lp(a) levels, 0.934 (0.90 to 0.97) for HDL-C and 0.96 (0.94 to 0.98) for Apolipoprotein A-I. The IVW odds ratio (95% CI) per doubling in the odds of genetically liability to binary risk factors was 2.05 (1.59 to 2.64) for hypertension, 1.20 (1.09 to 1.31) for nephrolithiasis and 1.08 (1.05 to 1.12) for T2D. The effects of these eight risk factors on CKD was consistent across UK biobank, CKDGen and HUNT (**Supplementary Table 7A, 8A** and **9A**).

In East Asian participants, genetic predicted level of BMI (OR = 1.42, 95%CI = 1.20 to 1.69, P = 6.49 × 10^−5^), increased nephrolithiasis risk (OR = 1.12, 95%CI = 1.04 to 1.19, P = 1.11 × 10^−3^) and increased T2D risk (OR = 1.07, 95%CI = 1.03 to 1.10, P = 1.66 × 10^−4^) were associated with increasing risk of CKD (**Supplementary Figure 2B**). The effects of three factors on CKD was consistent across three East Asian studies (except that for BMI and nephrolithiasis effect on CKD was not observed in the China Kadoorie Biobank, which had limited CKD cases) (**Supplementary Table 10, 11** and **12**).

For sensitivity MR analyses, bidirectional MR analysis found a consistent effect of increased genetic liability of CKD on increasing hypertension in European ancestry (**Supplementary Table 13**). The MVMR results of T2D, BMI and hypertension on CKD can be found in **Supplementary Table 14 A, B** and **C**. MR analysis using eGFR as outcome showed similar results as CKD (**Supplementary Table 7B, 8B, 9B** and **10B**).

#### Risk factors showing weak MR evidence

For the remaining 37 risk factors in European ancestry (**Supplementary Table 7, 8** and **9**) and 12 risk factors in East Asian ancestry (**Supplementary Table 10, 11** and **12**), weak evidence was found to support their effect on CKD. This included some established risk factors such as smoking and serum uric acid. In addition, shorter sleep duration showed evidence of association with CKD in Japan-Kidney-Biobank/ToMMo (**Supplementary Table 11**) and in the UK Biobank (**Supplementary Table 7A**) but lack of replication in other studies.

### Follow-up MR analyses of key findings

#### Effects of glycemic phenotypes and CKD

Although the evidence for an effect of T2D on CKD was reliable, we detected little evidence of the effect of eight glycemic phenotypes (fasting insulin (FI), fasting glucose (FG), 2-hour glucose (2hGlu), fasting proinsulin (FP), haemoglobin A1c (HbA1c), HOMA-B, Insulin-Like Growth Factor Binding Protein 3 and Insulin-Like Growth Factor I) on CKD (**Supplementary Figure 3**) and eGFR (**Supplementary Figure 4**). To further evaluated these findings, we found that 1) similar results were observed after controlling for possible reverse causation of instruments and potential outliers (**Supplementary Table 15**); 2) little evidence was observed that genetic liability to T1D was associated with CKD risk in any of the three outcome studies from European ancestry (**Supplementary Table 16A**), which further supported the weak effect of glucose on CKD; 3) for the MR analysis using stratified eGFR in Europeans, little effect of glycemic phenotypes on eGFR was observed in both diabetic and non-diabetic samples (**Supplementary Table 16B**), which suggested that the weak effect of glucose on CKD could be independent to diabetes; 4) fasting glucose and genetic liability to T2D were associated with diabetic retinopathy (**Supplementary Table 16C**), suggesting that the genetic predictors of glycemic phenotypes used for the main MR analyses were reliable.

#### Effects of blood lipids and CKD

For the MR findings of lipids, our follow-up analyses showed a few novel observations: 1) we observed different MR evidence for HDL-C on CKD across Europeans and East Asians. In Europeans, good MR strength were observed to support the effects of HDL-C and apolipoprotein A-I on CKD (**Supplementary Table 7, 8** and **9**), while HDL-C showed weaker MR strength in East Asians (**Supplementary Table 10, 11** and **12**). To test the potential influenced of power of the HDL-C effect on CKD in East Asians (OR = 0.94, 95%CI = 0.87 to 1.02), we conducted MR using better powered HDL-C associated SNPs in Europeans as genetic predictors (but the genetic associations were still from East Asian studies) and found reliable MR evidence between the two (OR = 0.89, 95%CI = 0.83 to 0.96; **Supplementary Table 17A**), which suggested HDL-C may has an effect on CKD in both population; 2) using European data, the multivariable MR model considering both HDL-C and apolipoprotein A-I in the same model showed that the effect of HDL-C on CKD were independent to the apolipoprotein A-I (**Supplementary Table 14D**); 3) following the HDL-C finding, we found an effect of circulating CETP level on CKD in CKDGen (OR = 1.06, 95%CI = 1.01 to 1.11, P = 0.01; **Supplementary Table 18**); 4) we further investigated the potential influence of apolipoprotein(a) size on CKD, but found little evidence for a causal effect. This suggested that the effect of Lp(a) level on CKD was independent of apolipoprotein(a) size (**Supplementary Table 19**);

#### Effect of blood pressure phenotypes and CKD across populations

As shown in **Figure 3**, blood pressure phenotypes including hypertension, SBP and PP showed reliable evidence of strong causal effects on CKD in the European studies but appeared to show a null causal effect in the East Asian studies (ORs of hypertension on CKD ranging from 1.72 to 2.35 in Europeans but only from 0.99 to 1.07 in East Asians). To validate the East Asian results, we first conducted a MR analysis using the more powerful European SBP and DBP instruments (but East Asian association information), which showed similar results (**Supplementary Table 17B**). Second, we used genetic instruments of SBP, DBP and hypertension from an additional GWAS study in East Asia (40) and still observed similar results (**Supplementary Table 17C**). These analyses provided additional evidence that blood pressure might have a population-specific role in CKD aetiology.

**Figure 3.**
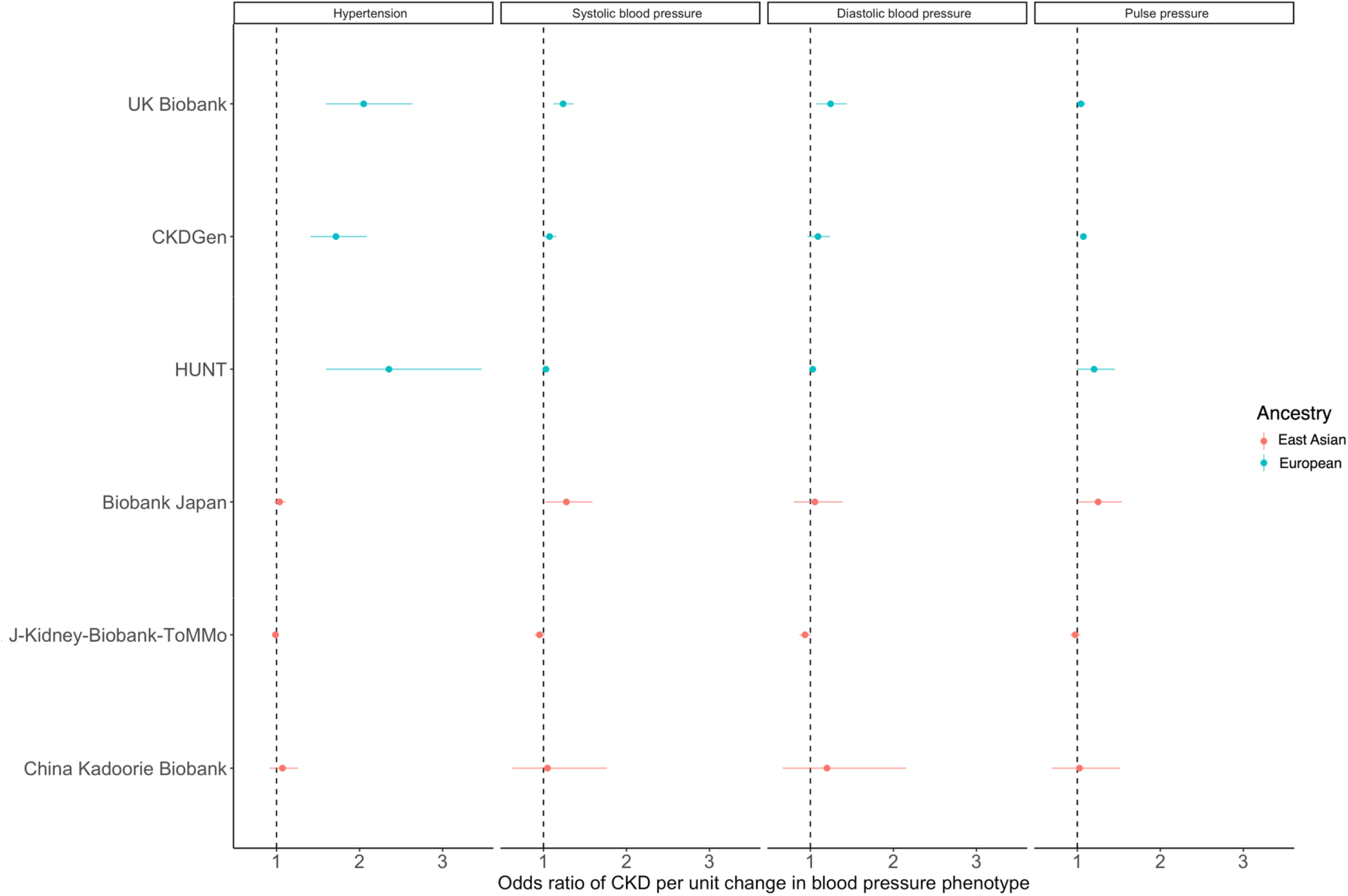
Forest plot for causal effects of four blood pressure phenotypes on CKD risk. The subplots represent MR results of different blood pressure phenotypes.

#### Non-linear effects of BMI and fasting glucose on CKD

We observed a threshold relationship between BMI and CKD. The curved shape of this relationship suggested higher risk in overweight or obese participants, with the BMI threshold at around 25 kg/m^2^ in UK Biobank and around 24∼25 kg/m^2^ in HUNT Study (**Figure 4**). Stratified analysis split by sex suggested similar effects of BMI on CKD (**Supplementary Figure 5**), while stratified analysis split by age showed greater harm of increasing BMI in older participants (age> = 65) (**Supplementary Figure 6**).

**Figure 4.**
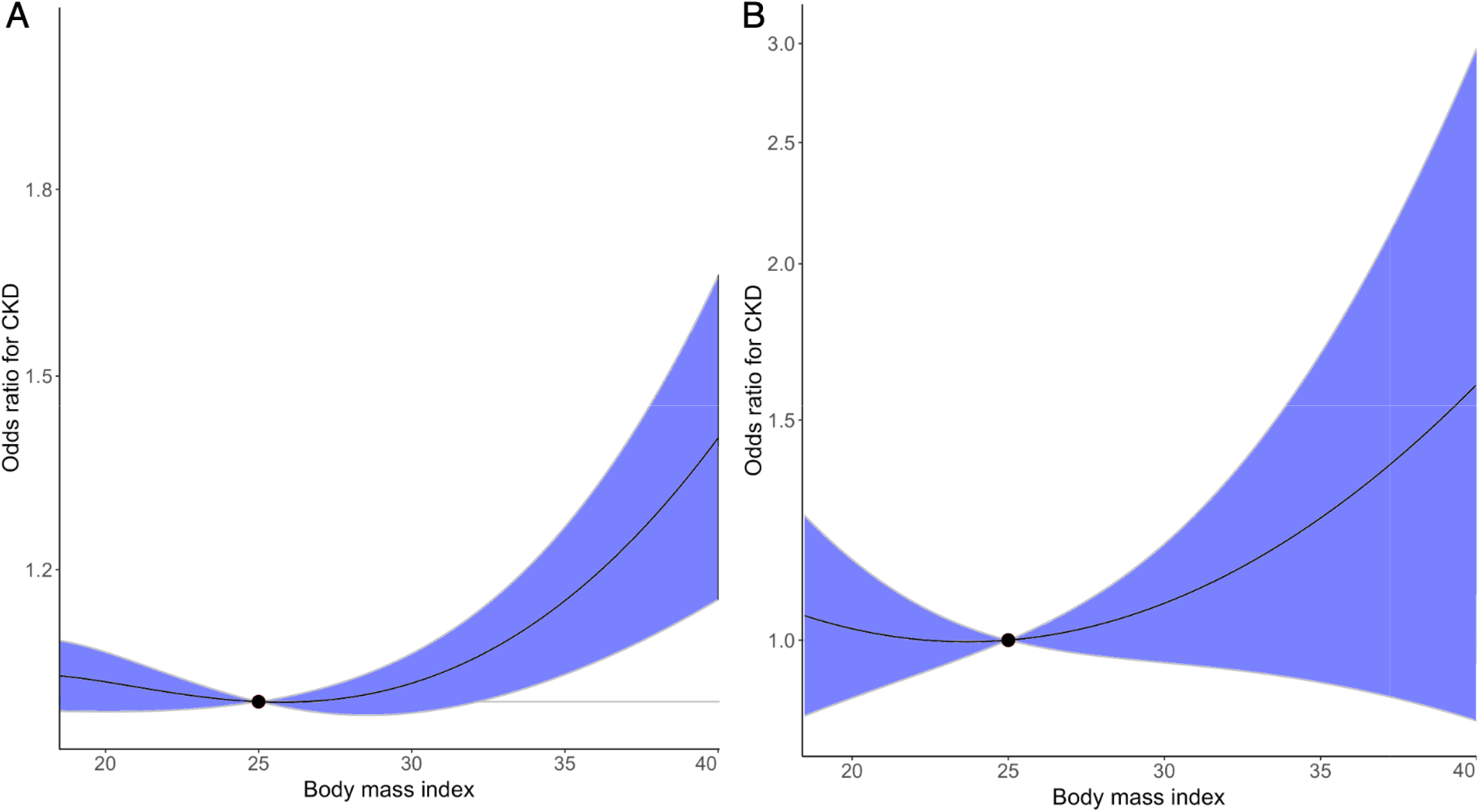
Non-linear Mendelian randomization of BMI on CKD risk. The dose-response curve between body mass index and CKD risk for (A) UK Biobank and (B) HUNT Study. Gradient at each point of the curve is the localized average causal effect. Shaded areas represent 95% confidence intervals.

Fasting glucose showed weak evidence for a non-linear relationship with CKD (**Supplementary Figure 7**). This finding was consistent with the stratified analysis results split by sex (**Supplementary Figure 8**) and age (**Supplementary Figure 9**). **Supplementary Table 20** showed the non-linear trend test results of BMI and fasting glucose on CKD, where the analyses suggested little evidence supporting a non-linear relationship between fasting glucose and CKD.

## DISCUSSION

In this trans-ethnic MR study, we comprehensively assessed the causal relationships of 45 reported risk factors on CKD and eGFR in more than 1.1 million European and 17 risk factors in over 250,000 East Asian ancestries. Using a set of MR approaches including five two-sample MR methods and multivariable MR, we found reliable evidence of causal effects of eight cardio-metabolic related risk factors (BMI, SBP, hypertension, T2D, nephrolithiasis, HDL-C, apolipoprotein A-I, and Lp[a]) on CKD. The remaining 37 risk factors, such as smoking and serum uric acid, showed weak evidence of causal effects on CKD with the current available data. These findings agreed with previous MR studies of similar risk factors separately (42)(43)(44). The null finding of the serum uric acid agreed with the recent trial evidence of the effects of serum urate lowering (using Allopurinol) on CKD progression (45)(46). Notably, our extensive MR and follow-up analyses suggested the possibility of glucose-independent pathways linking T2D with CKD. Using non-linear MR, we observed a threshold relation between BMI and CKD risk, with the CKD risk increased at BMI above 25 kg/m^2^.

The causal patterns of 17 risk factors were compared across the two ancestries, and we observed consistent effects of T2D, BMI and nephrolithiasis on CKD in both Europeans and East Asians. In contrast, distinguishable causal patterns were observed for hypertension against CKD across ancestries, where the causal estimate was strong in Europeans but towards null in East Asians. Our trans-ethnic comparison implies that careful assessments are needed before implementing interventions using CKD risk factors that were identified using evidence from a different population.

For the prioritised risk factors, hypertension is one of the most common risk factors for CKD progression (51). A recent bi-directional MR study in Europeans supported the causal effects of higher kidney function on lower blood pressure using well-designed eGFR instruments controlled by blood urea nitrogen. But the study suggested inconclusive evidence of an effect of blood pressure on eGFR (52). In our MR analysis, we found evidence of positive bidirectional causal effects between hypertension and CKD in Europeans, in line with clinical and experimental studies. The inconsistent MR findings across these studies may be due to 1) the blood pressure instruments Yu et al used were adjusted for BMI in the additive model, but the genetic associations of eGFR and CKD were not controlled for BMI. Given the causal role of BMI on both CKD and hypertension, only controlling for BMI in the exposure data may create unintended bias to the MR estimates, as described previously (53); 2) the CKD cases we used here were clinically diagnosed, which may bring additional statistical power and therefore provided more reliable evidence for the suggestive effect of blood pressure on CKD reported by Yu et al. Furthermore, the different MR evidence across ancestries we observed combined with previous literature evidence opens up the possibility that hypertension could play different causal roles on CKD across ancestries. In literature, the ethnic disparities for effects of hypertension on CKD has been reported (47)(48). For example, Chinese hold lower risk of CKD when exposed to hypertension compared with Europeans (47). In 2019, hypertensive nephropathy accounted for 27% of the overall CKD cases in U.S but only accounted for 20.8% in East Asian (49)(50). Correctively, whether blood pressure showed a consistent causal effect on CKD across ancestries is still not directly clear. Well-powered trans-ethnic studies are needed to validate our findings.

For two other emerging risk factors, BMI and nephrolithiasis, our MR analyses suggested substantial causal effects on CKD. Previous observational studies suggested a positive association for BMI on CKD (54)(55) and end stage renal disease (ESRD) onset (56) (57)(58) and the effect of weight loss on slowing kidney function decline (59). Using linear and nonlinear MR approaches, we observed a threshold causal relationship between BMI and CKD and quantified the optimal maximum (25 kg/m^2^) for body mass index. These findings provide additional confidence of the potential success of five ongoing trials registered in http://clincaltrials.gov. In addition, nephrolithiasis is a common serious problem around the world (60)(61)(62). Increasing evidence suggests that having kidney stones is a risk factor for CKD (63). For example, kidney stone formers tend to have lower eGFR (64). Previous cohort study suggested that even a single kidney stone episode was associated with an increase in the likelihood of adverse renal outcomes (65). A recent genetic study suggested that eGFR has an inverse relationship with kidney stone formation (66). However, the causal relationship between nephrolithiasis and CKD has not been investigated thoroughly. Our MR supported the causal effect of nephrolithiasis on increased risk of CKD, which were obtained by genetic predictors with good instrument strength. This is of particular importance as obstructive nephropathy was the third leading cause of CKD in the general Chinese population (15.6%) (50). Existing literature suggests that recurrent obstructive episodes may activate the fibrogenic cascade in renal cells and be responsible for the loss of renal function (67). Our study supports this literature and prioritises prevention of kidney stones as a sensible approach for CKD prevention.

Notably, diabetic kidney disease is considered the most common type of CKD worldwide(68). A previous MR study of T2D on CKD in Chinese suggested a strong causal link between the two (69), which aligned with our MR finding in East Asians and Europeans. However, despite the reliable evidence for a causal effect of T2D on CKD, the linear and non-linear MR results suggested that the causal effects of fasting glucose and insulin levels on CKD were close to null. It has been observed that with increasing use of glucose-lowering medications, the prevalence of CKD in diabetics was not reduced as expected from 1988 to 2008 (70). Meta-analysis of RCTs of intensive glucose control showed little effect on reducing the risk of ESRD during the years of follow-up (71). These findings together with our MR results suggest that non-glucose pathways could play a role to link diabetes with CKD (70)(71). Consistently, the beneficial effects of antidiabetic SGLT2 inhibitors on renal outcomes were suggested to be mediated by glucose-independent pathways (72). One potential limitation was that the glucose GWAS used in this study was conducted in a general population sample (fasting glucose level lower than 7 mmol/L), which most of the existing MR studies using these data assume the glucose change in general population shows similar effects in diabetes patients (with fasting glucose level above 7 mmol/L), which may not always be the case. Although our stratified MR analysis showed similar little causal effects using eGFR in diabetes patients and non-diabetes patients, we believe better genetic instruments from diabetic population samples and well-designed clinical trials are needed to evaluate the effect of glucose-dependent and –independent mechanisms on CKD prevention in diabetic patients.

Hyperlipidaemia and dyslipidaemia have been widely documented to be associated with kidney disease (73)(74). But the causal effects of lipid components on CKD are still unclear. A few recent MR studies suggested a protective effect of higher HDL-C on CKD in Europeans (75) and a nominal effect of Lp(a) lowering on reducing CKD risk (76). In this study, we strengthened the evidence of the Lp(a) finding and validated the HDL-C findings in completely independent samples. Our study also moved beyond the existing studies, by 1) establishing novel causal evidence of apolipoprotein A-I on CKD in Europeans; 2) the MVMR implies that non-apolipoprotein A-I property of HDL-C may have an effect on CKD; 3) providing a natural extension of the HDL-C finding (77) and a recent trial of CETP inhibitor on cardiovascular disease (78), by estimating the causal effect of circulating CETP level on CKD and demonstrating a suggested effect between the two in European ancestry; 4) demonstrating that the causal effect of Lp(a) level on CKD was independent of apolipoprotein(a) size. A previous study suggested that HDL-C may protect from kidney diseases through altering cholesterol efflux in the pathogenesis of kidney disease (79). The causal effect of HDL-C and the effect of CETP level on CKD raises the possibility of increasing HDL-C concentration as an intervention strategy for CKD prevention. In addition, our MR finding of the effects of Lp(a) level also aligned with previous observational evidence, which suggested that Lp(a) level was inversely correlated with kidney function (80)(81) and positively correlated with lower eGFR (82). This also implies the possibility of further investigation of Lp(a) reduction therapies, such as IONISAPO(a) on reducing CKD risk (83). Overall, our findings highlight the potential of multiple lipid management strategies in reducing CKD risk.

### Strengths and Limitations

Our study has some strengths compared to previous studies. We used clinically diagnosed CKD instead of eGFR < 60ml/min/1.73m^2^ in European (UK Biobank and HUNT) and East Asian studies (China Kadoorie Biobank and Japan-Kidney-Biobank/ToMMo), which the later definition excluded CKD cases with abnormal urine protein level but with normal eGFR. Therefore, this increased the robustness of the CKD definition. By comprehensively validating the MR findings in the six CKD studies, we greatly enhanced the reliability of the causal atlas of risk factors on CKD.

Our study also has some potential limitations. In the MR analysis, when instrumenting for a binary exposure (e.g. coronary artery disease), we are not instrumenting the exposure itself but the predisposition to the exposure (84), so our results speak to whether removal of the predisposition to the binary exposure (rather than treatment of the exposure) would reduce CKD risk. In addition, due to unbalanced representativeness of GWAS samples across Europeans and East Asians, we can only test causal effects for 17 of the 45 risk factors in East Asians. Due to the same reason, the number of instruments for each risk factors of the analyses were different across the two ancestries. For risk factors showed different MR evidence across ancestries, we further evaluated these findings using more powerful genetic predictors and using additional data sources to minimise the influence of unbalanced GWAS samples and number of instruments across ancestries. Other limitations of the study were listed in **Supplementary Note 6**.

### Conclusions

By evaluating causal evidence of 45 risk factors on CKD in over 1.1 million individuals in European ancestry and 17 factors in over 250,000 East Asian ancestry, we built up a causal atlas of CKD risk factors and showed that eight risk factors are reliably causal for CKD. All these factors are predominantly related to cardio-metabolic health, which supports the shared causal link between cardio-metabolic health and kidney function. Our findings may have important clinical implications in terms of informing primary prevention in “at risk” individuals with normal renal function, which may help reduce the burden of CKD globally.

## Data Availability

The genome-wide association analysis results of CKD and eGFR in the UK Biobank can be accessed via MR-Base (www.mrbase.org) and the IEU OpenGWAS database (gwas.mrcieu.ac.uk).

https://gwas.mrcieu.ac.uk

## Author contribution

J.Z., Y.M.Z. and H.R. performed the linear Mendelian randomization analysis; J.Z. and H.R. performed the non-linear Mendelian randomization with supports from S.F., Q.Y. and S.B.; J.Z., H.R. and L.F.T. performed the GWAS in the UK Biobank and HUNT study; Y.S., M.Y. and N.K. conducted the GWAS in each cohort and performed the GWAS meta-analysis in the Japan-Kidney-Biobank/ToMMo study; M.A., M.K., K.M., Y.K., and Y.O. performed the GWAS in Biobank Japan; C.Q.Y. and J.C.L conducted the GWAS in the China Kadoorie Biobank; J.Z., Y.M.Z and B.E. performed the systematic review of CKD risk factors; R.E.W. performed the sensitivity analysis of smoking on CKD; P.C.H., A.H., J.R., B.M.B., L.F.T., K.H., S.H. A.K., C.P., M.W., and B.O.A. provided key data and supported the MR analysis; M.C.B., Y.C., R.C., S.H., N.F., A.P.M., G.D.S., S.B., C.Q.Y. and B.O.A. reviewed the paper and provided key comments; J.Z., Y.M.Z., H.R., V.W., Y.S., Y.L., X.M., Y.F.B., G.N., G.D.S., S.B., B.O.A, H.Z. and T.R.G. wrote the manuscript; J.Z., Y.M.Z., H.Z. and T.R.G. conceived and designed the study and oversaw all analyses.

## Acknowledgements

JZ is funded by a Vice-Chancellor Fellowship from the University of Bristol. This research was also funded by the UK Medical Research Council Integrative Epidemiology Unit (MC_UU_00011/1, MC_UU_00011/4 and MC_UU_00011/7). This study was funded/supported by the NIHR Biomedical Research Centre at University Hospitals Bristol NHS Foundation Trust and the University of Bristol (GDS, TRG and REW). This study received funding from the UK Medical Research Council (MR/R013942/1). The views expressed in this publication are those of the author(s) and not necessarily those of the NHS, the National Institute for Health Research or the Department of Health and Social Care. YMZ is supported by the National Natural Science Foundation of China (81800636) and Beijing Natural Science Foundation (7184253). HZ is supported by the Training Program of the Major Research Plan of the National Natural Science Foundation of China (91642120), the Grant from the Science and Technology Project of Beijing, China (D18110700010000) and the University of Michigan Health System–Peking University Health Science Center Joint Institute for Translational and Clinical Research (BMU2017JI007). NF is supported by the National Institutes of Health awards R01-MD012765, R01-DK117445 and R21-HL140385. RC is funded by a Wellcome Trust GW4 Clinical Academic Training Fellowship (WT 212557/Z/18/Z). The Nord-Trøndelag Health Study (The HUNT Study) is a collaboration between HUNT Research Centre (Faculty of Medicine and Health Sciences, NTNU, Norwegian University of Science and Technology), Nord-Trøndelag County Council, Central Norway Regional Health Authority, and the Norwegian Institute of Public Health. MCB is supported by the UK Medical Research Council (MRC) Skills Development Fellowship (MR/P014054/1). SF is supported by a Wellcome Trust PhD studentship (WT108902/Z/15/Z). QY is funded by a China Scholarship Council PhD scholarship (CSC201808060273). YC was supported by the National Key R&D Program of China (2016YFC0900500, 2016YFC0900501, 2016YFC0900504). The China Kadoorie Biobank baseline survey and the first re-survey were supported by a grant from the Kadoorie Charitable Foundation in Hong Kong. The long-term follow-up is supported by grants from the UK Wellcome Trust (202922/Z/16/Z, 088158/Z/09/Z, 104085/Z/14/Z). Japan-Kidney-Biobank was supported by AMED under Grant Number 20km0405210. PCH is supported by Cancer Research UK [grant number: C18281/A19169]. AK was supported by DFG KO 3598/5–1. N.F. is supported by NIH awards R01-DK117445, R01-MD012765, R21-HL140385.

